# Prevalence and Determinants of Scabies Infection among Male Inmates at Kiambu Main Prison, Kenya: A Mixed-Methods Cross-Sectional Study

**DOI:** 10.64898/2026.08.03.26359545

**Authors:** Esther Makokha Kulundu, Wycliffe Nyamongo Onkoba, Chrispine Ochieng Ngwawe

**Author notes:** Corresponding author, Contact details, Personal.

## Abstract

**Background:** Scabies remains a major neglected tropical disease in correctional facilities due to overcrowding and limited hygiene resources. However, evidence regarding the determinants of scabies infection among inmates in Kenyan prisons remains limited. This study assessed the prevalence and determinants of scabies infection among male inmates at Kiambu Main Prison, Kiambu County, Kenya.

**Methodology:** A mixed-methods analytical cross-sectional study was conducted among 188 male inmates recruited using a ward-stratified pragmatic sampling approach between May and June, 2026. Quantitative data were collected using questionnaires and clinical examinations based on the International Alliance for the Control of Scabies (IACS) diagnostic criteria. Qualitative data were obtained through focus group discussions and key informant interviews. Quantitative data were analysed using descriptive statistics, chi-square tests, and logistic regression, whereas qualitative data were analysed thematically.

**Results:** The prevalence of scabies was 54.8%, with a weighted prevalence of 54.0% (95% CI: 46.5– 61.6%). Daily bathing (AOR = 0.07, 95% CI: 0.01–0.31), bathing 2-3 times per week (AOR = 0.09, 95% CI: 0.01–0.44), and daily clothes washing (AOR = 0.21, 95% CI: 0.06–0.65) were independently associated with lower odds of scabies infection. Remandees had higher odds of infection than convicted prisoners (AOR = 1.97, 95% CI: 1.00–3.91). Qualitative findings identified overcrowding, inadequate hygiene resources, limited health education, medication shortage, and delays in treatment as key contributors to scabies transmission.

**Conclusion:** Scabies remains highly prevalent among inmates at Kiambu Main Prison. Both individual hygiene practices and institutional conditions contribute to disease transmission. Strengthening prison health services through routine screening, structured health education, improved access to hygiene resources, and timely diagnosis and treatment may help reduce transmission. These findings provide evidence to inform prison health policies and targeted scabies control programmes in Kenya and other resource-limited correctional settings.

**Author Summary:** Scabies is a highly contagious neglected tropical disease that disproportionately affects people living in overcrowded settings with limited access to water, sanitation, and healthcare, including prisons. Although incarcerated populations are particularly vulnerable to scabies transmission, little is known about the factors that contribute to its spread in Kenyan correctional facilities. We investigated the prevalence of scabies among male inmates at Kiambu Main Prison and explored the individual and institutional determinants of infection using questionnaires, clinical examinations, group discussions, and interviews with prison staff. More than half of the participants were found to have scabies. Regular bathing and daily washing of clothes were associated with a lower likelihood of scabies infection, while remandees were at greater risk than convicted prisoners. Inmates and prison staff also identified overcrowding, shortages of hygiene supplies and medicines, limited health education, and delays in diagnosis and treatment contributed to continued transmission. Our findings highlight practical opportunities to reduce the burden of scabies through routine screening, improved hygiene resources, health education, and timely treatment in prisons.

## Introduction

Scabies is a contagious parasitic skin disease caused by the mite *Sarcoptes scabiei var. hominis*, which burrows into the epidermis and deposits eggs, triggering an immune response that results in intense itching and rash (1,2). The disease is transmitted primarily through prolonged person-to-person (skin-to-skin) contact, with occasional indirect transmission through contaminated fomites such as clothing and bedding, particularly in cases of crusted scabies, and is closely associated with overcrowding, poor hygiene, and limited access to healthcare. In 2017, the World Health Organization (WHO) recognized scabies as a neglected tropical disease (NTD) because of the substantial health burden it imposes, particularly among socially and economically disadvantaged populations living in overcrowded conditions with limited access to healthcare, such as people in prisons, refugee camps, and underserved communities (3). Globally, approximately 200 million people are affected at any given time, with an estimated 450 million new cases occurring annually (2,4). Although scabies is rarely fatal, persistent infestation can lead to secondary bacterial infections, sleep disturbance, reduced quality of life, and, in severe cases, crusted scabies among immunocompromised individuals (5,6).

Inmates and remandees in prisons disproportionately bear the burden of scabies infection due to overcrowding, frequent close contact, inadequate hygiene and sanitation resources, and limited access to healthcare services. Delie et al. reported a pooled scabies prevalence of 19.6% among prisoners in Africa, substantially higher than the estimated global prevalence of 6.6% (2). These findings are supported by studies conducted by Gupta et al., Melese et al., and Bogino et al., which identified both individual hygiene practices and institutional conditions, including lack of soap use, infrequent bathing, sharing of clothing and bedding, poor ventilation, inadequate water supply, and overcrowded living conditions, as key drivers of scabies transmission in correctional facilities and other high-risk settings (6–8). For instance, scabies outbreaks have been reported in Kakamega, Busia, and Kisii prisons in Kenya in Kenya (9,10). However, evidence regarding the determinants of scabies infection among inmates in Kenyan prisons remains limited. Though there is association of such outbreaks to overcrowding, where Kenyan correctional facilities are housing approximately 58,000 inmates in facilities originally designed for about 34,000 individuals (11). Such overcrowding creates conditions that favour the rapid transmission of communicable skin diseases, including scabies. Routine clinical services at Kiambu Main Prison have documented recurrent scabies cases, yet no published epidemiological data are available to quantify the burden of disease or identify associated risk factors of transmission of scabies. Thus, the absence of prison-specific epidemiological evidence limits the development of targeted scabies prevention and control programmes in Kenyan prisons. Accordingly, this study aimed to determine the prevalence of scabies among male inmates at Kiambu Main Prison, identify the individual and institutional determinants associated with infection, and use qualitative evidence to provide contextual explanations for the quantitative findings. The findings provide evidence to inform targeted scabies prevention and control strategies in Kenyan prisons.

## Methods

### Ethics Statement

This study was conducted in accordance with the ethical principles of the Declaration of Helsinki. Ethical clearance was obtained from the AMREF Health Africa Ethics and Scientific Review Committee (Approval No. P2079/2026). Research authorization was granted by the National Commission for Science, Technology, and Innovation (NACOSTI), and permission to conduct the study was obtained from the Kenya Prisons Service Headquarters and the management of Kiambu Main Prison. Written informed consent was obtained from all participants prior to data collection and clinical examination. Participation was voluntary, and participants were informed of their right to decline or withdraw from the study at any stage without consequences. Confidentiality was maintained by assigning unique identification codes instead of participant names, and all study records were stored securely with access limited to the research team. Participants diagnosed with scabies during the clinical examination were treated with benzyl benzoate.

The findings are reported in accordance with the Strengthening the Reporting of Observational Studies in Epidemiology (STROBE) guidelines.

### Study Design and Setting

This study employed a facility-based convergent mixed-methods analytical cross-sectional design among male inmates at Kiambu Main Prison, Kiambu County, Kenya. The quantitative component involved structured questionnaires and clinical examinations for scabies, while the qualitative component comprised focus group discussions (FGDs) with inmates and key informant interviews (KIIs) with prison staff to contextualize the quantitative findings. Kiambu Main Prison, located in Kiambu County, Kenya, is a correctional facility for male inmates. During the study period (May- June 2026), the prison housed 732 inmates despite an official capacity of approximately 450, representing an occupancy level of approximately 163%. Inmates reside in shared wards and have frequent close interpersonal contact, conditions that facilitate the transmission of communicable skin diseases such as scabies. The prison has an on-site health clinic that provides routine healthcare services, including outpatient consultations, diagnosis, and treatment of common illnesses.

### Study Population

The study population comprised male inmates incarcerated at Kiambu Main Prison during the study period. Eligible participants were inmates aged 18 years or older who had been incarcerated for at least three weeks to allow for the incubation period of scabies (12) and who provided written informed consent. Inmates with dermatological conditions likely to interfere with clinical diagnosis, those who were critically ill or physically unable to participate, and those receiving treatment for other infectious diseases during the study period were excluded.

### Sample Size and Sampling Procedures

The sample size was determined using Cochran’s formula (1977) for prevalence studies, assuming a 95% confidence level, 5% margin of error, and an estimated scabies prevalence of 20% derived from the pooled prevalence reported by Delie et al. (2024) in their systematic review and meta-analysis of scabies among prisoners in Africa (2). After applying the finite population correction for the 732 inmates, the minimum required sample size was 185. A total of 189 eligible inmates were recruited to account for potential non-response. One participant withdrew before completing the questionnaire and was excluded from the analysis, resulting in a final analytical sample of 188 inmates. The prison population consisted of two administrative strata: convicted prisoners (n = 200) and remandees (n = 532). The required sample was initially allocated proportionately across prison wards according to the number of inmates in each ward. Within each ward, simple random sampling using inmate registers was planned. During implementation, however, operational constraints inherent to the correctional setting, including court appearances, visitation schedules, and the requirement for voluntary informed consent, limited the feasibility of strict random selection. Consequently, participant recruitment followed a ward-stratified pragmatic sampling approach, whereby eligible inmates who were available and consented to participate during data collection were recruited while maintaining the planned proportional allocation wherever feasible. The final sample comprised 49 convicted prisoners and 139 remandees.

During the study period, Ward 10 served as a temporary isolation ward for inmates with confirmed or suspected scabies who had been transferred from other prison wards for clinical management. To ensure adequate representation of clinically confirmed scabies cases, additional eligible inmates from this ward were recruited. Because this resulted in unequal sampling fractions across wards, ward-specific sampling weights, calculated as the inverse of the sampling fraction within each ward (ward population divided by the number of inmates sampled), were applied during prevalence estimation to generate estimates representative of the overall prison population. Logistic regression analyses were performed using the observed sample without weighting.

Purposive sampling was used to recruit participants for the qualitative component. Two FGDs, each comprising eight inmates, were conducted to explore experiences related to hygiene practices, living conditions, access to water, and healthcare-seeking behaviour. Participants were stratified by duration of incarceration (<3 months and ≥3 months), with both groups including convicted prisoners and remandees. Four key informant interviews (KIIs) were conducted with the prison clinical officer, nursing officer, welfare officer, and a senior prison warden. Interviews continued until thematic saturation was achieved.

### Data Collection Procedures

Data were collected between 19^th^ May and 5^th^ June 2026 using quantitative and qualitative methods. The questionnaire was pretested among inmate trustees who were not included in the final study to assess clarity, relevance, and feasibility.

Quantitative data were collected using self-administered structured questionnaires and clinical examinations for scabies. The questionnaire captured participants’ socio-demographic, incarceration, hygiene, and institutional characteristics. Questionnaires were completed independently by participants, with assistance provided to those who had difficulty reading or understanding the questions. Scabies diagnosis was conducted by trained healthcare personnel using the International Alliance for the Control of Scabies (IACS) diagnostic criteria (13). FGDs and KIIs were conducted using semi-structured interview guides. Discussions and interviews were audio-recorded with participants’ consent, supplemented by field notes, and conducted in private locations within the prison to ensure confidentiality while maintaining institutional security.

The primary outcome was scabies infection, classified as either present or absent based on clinical diagnosis using the IACS diagnostic criteria. Independent variables included individual factors (age, education, bathing frequency, soap use, washing of clothes and bedding, and sharing of personal items) and institutional factors (prison status, duration of imprisonment, ward occupancy level, the number of inmates sharing sleeping spaces, and receipt of health education on scabies, and reported lack of medication for scabies treatment). Health education and medication availability were included as proxy indicators of institutional prevention and treatment capacity.

Ward occupancy level was defined as the number of inmates housed in each prison ward during the study period. For analysis, ward occupancy was categorised into four levels (low, moderate, high, and very high) using cut-off values of ≤44, 45–61, 62–82, and >82 inmates, respectively. These categories were based on the distribution of ward occupancy within the study population, with cut-off points corresponding approximately to the quartiles of the occupancy distribution to ensure adequate numbers of participants in each category for statistical comparison.

Most explanatory variables were analysed as categorical variables. Age was grouped into 18–29, 30–39, 40–49, and ≥50 years, while duration of incarceration was categorised as ≤3 months and >3 months. Hygiene-related variables, including bathing frequency, clothes washing frequency, and bedding washing frequency, were analysed according to their original questionnaire response categories. Prison status, health education, medication availability, and sharing of personal items were analysed as binary variables. Categories were defined a priori based on the questionnaire response options and their epidemiological relevance to facilitate meaningful comparisons in the logistic regression analyses.

Several measures were implemented to minimise bias and improve data quality. A stratified sampling approach was used to enhance the representativeness of the study population and reduce selection bias. Sampling weights were applied during prevalence estimation to account for the disproportionate sampling of participants from Ward 10 and to produce estimates representative of the overall prison population. The questionnaire was pretested before the main study to assess its clarity, relevance, and feasibility, thereby reducing measurement error during data collection. In addition, scabies diagnosis was based on the International Alliance for the Control of Scabies (IACS) diagnostic criteria, applied by trained healthcare personnel to ensure consistent and accurate case identification.

### Data Analysis

Quantitative data were analysed using R Statistical Software (version 4.5.3). Descriptive statistics were used to summarize participant characteristics and study variables. Weighted prevalence estimates and 95% confidence intervals (CIs) were generated using survey analysis procedures to account for disproportionate sampling across prison wards. Sampling weights were applied only for estimating prevalence because oversampling was designed to improve precision of prevalence estimates rather than produce weighted exposure-outcome associations. Consequently, logistic regression analyses were performed using the observed (unweighted) sample.

Bivariate associations between scabies infection and individual and institutional factors were assessed using Pearson’s chi-square test or Fisher’s exact test, as appropriate. Univariable logistic regression was used to estimate crude odds ratios (CORs). Variables with a p-value < 0.20 in the bivariate analysis, together with variables considered biologically plausible based on existing literature, were included in multivariable logistic regression models to identify independent determinants of scabies infection. Separate multivariable models were fitted for individual and institutional factors. Adjusted odds ratios (AORs) with 95% confidence intervals were reported, and statistical significance was set at p < 0.05. Final models were assessed for multicollinearity using the Generalised Variance Inflation Factor (GVIF) and for goodness-of-fit using the Hosmer–Lemeshow test. Qualitative data from focus group discussions and key informant interviews were transcribed verbatim and analysed thematically using NVivo (version 15). Themes were developed deductively from the study objectives and inductively from participants’ narratives, with representative quotations used to provide contextual explanations for the quantitative findings.

## Results

### Participant Recruitment

A total of 189 male inmates were recruited into the quantitative study. One participant withdrew before completing the questionnaire and was therefore excluded from analysis, resulting in a final analytical sample of 188 inmates (Fig 1).

**Fig 1:**
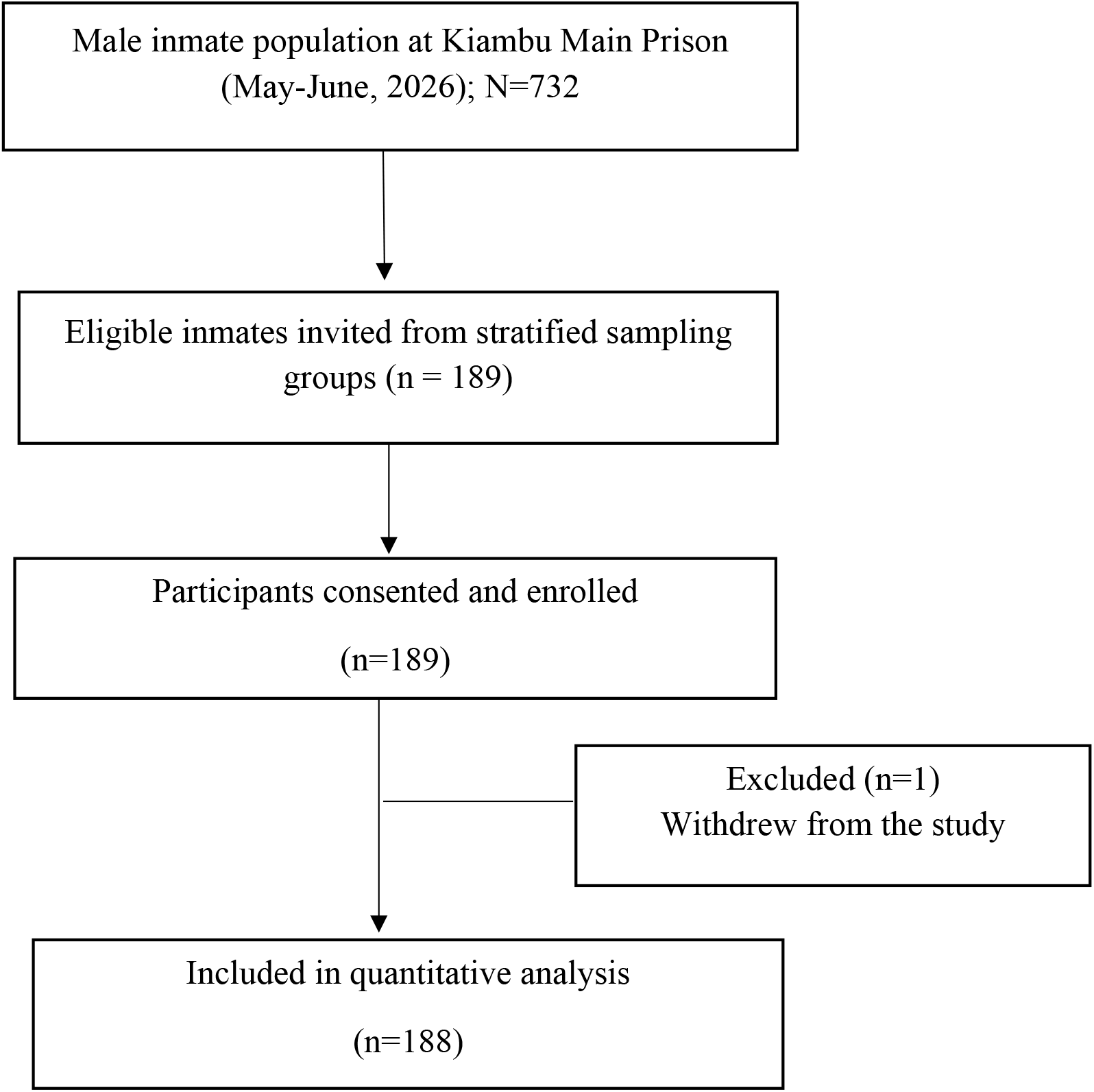
Flow diagram of participant recruitment and inclusion in the quantitative analysis.

### Participant Characteristics

Among the analysed participants, most were aged 18–39 years (77.2%) and had attained primary (41.0%) or secondary (45.7%) education. Nearly three-quarters (73.9%) were remandees, two-thirds (66.5%) had been incarcerated for more than three months, and 69.1% reported sharing sleeping space with more than two inmates (Table 1).

**Table 1:** Characteristics of the study participants (N = 188)

| Characteristic | n (%) |
| --- | --- |
| **Age group (years)** |  |
| 18–29 | 80 (42.6) |
| 30–39 | 65 (34.6) |
| 40–49 | 30 (16.0) |
| ≥50 | 13 (6.9) |
| **Education level** |  |
| None | 5 (2.7) |
| Primary | 77 (41.0) |
| Secondary | 86 (45.7) |
| College or higher | 20 (10.6) |
| **Prison status** |  |
| Convicted prisoner | 49 (26.1) |
| Remandee | 139 (73.9) |
| **Duration of incarceration** |  |
| ≤3 months | 63 (33.5) |
| >3 months | 125 (66.5) |
| **Number sharing sleeping space** |  |
| ≤2 inmates | 58 (30.9) |
| >2 inmates | 130 (69.1) |

Values are presented as *n* (%).

### Prevalence

Of the 188 participants, 103 were clinically diagnosed with scabies, giving a crude prevalence of 54.8% (95% CI: 47.6–61.9%). After adjustment for the stratified sampling design, the weighted prevalence was 54.0% (95% CI: 46.5–61.6%) (Fig 2).

**Fig 2:**
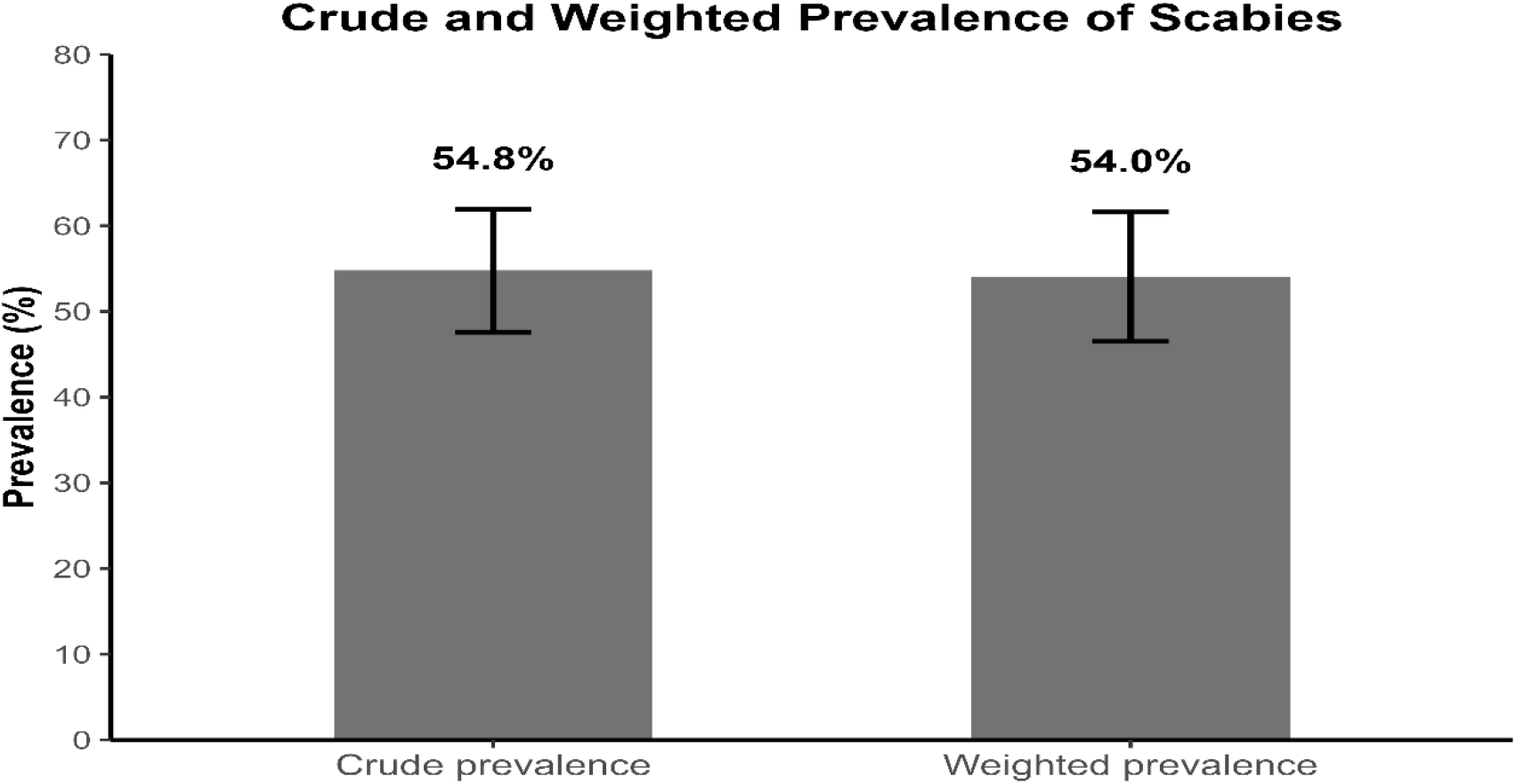
Crude and weighted prevalence of scabies among male inmates at Kiambu Main Prison.

### Factors associated with scabies infection

Hygiene-related practices differed significantly according to scabies status. Compared with inmates without scabies, infected inmates reported less frequent bathing, clothes washing, and bedding washing (Table 2). Age, education, duration of incarceration, ward occupancy level, soap use, and sharing of sleeping space were not significantly associated with infection.

**Table 2:** Socio-demographic, institutional and hygiene characteristics of inmates by scabies infection status.

| Characteristic | No<br>N = 85<sup>1</sup> | Yes<br>N = 103<sup>1</sup> | p-value<sup>2</sup> |
| --- | --- | --- | --- |
| **Age group (years)** |  |  | 0.5 |
| 18–29 | 35 (41.2%) | 45 (43.7%) |  |
| 30–39 | 31 (36.5%) | 34 (33.0%) |  |
| 40–49 | 11 (12.9%) | 19 (18.4%) |  |
| 50+ | 8 (9.4%) | 5 (4.9%) |  |
| **Education level** |  |  | 0.3 |
| None | 1 (1.2%) | 4 (3.9%) |  |
| Primary | 37 (43.5%) | 40 (38.8%) |  |
| Secondary | 41 (48.2%) | 45 (43.7%) |  |
| College+ | 6 (7.1%) | 14 (13.6%) |  |
| **Prison status** |  |  | 0.11 |
| Prisoner | 27 (32%) | 22 (21%) |  |
| Remandee | 58 (68%) | 81 (79%) |  |
| **Duration of incarceration** |  |  | 0.2 |
| ≤3 months | 33 (38.8%) | 30 (29.1%) |  |
| >3 months | 52 (61.2%) | 73 (70.9%) |  |
| **Ward occupancy level** |  |  | 0.8 |
| Low | 21 (25%) | 28 (27%) |  |
| Moderate | 26 (31%) | 31 (30%) |  |
| High | 28 (33%) | 36 (35%) |  |
| Very High | 10 (12%) | 8 (7.8%) |  |
| **Number sharing sleeping space** |  |  | 0.6 |
| ≤2 | 28 (33%) | 30 (29%) |  |
| >2 | 57 (67%) | 73 (71%) |  |
| **Bathing frequency** |  |  | <0.001 |
| Rarely | 2 (2.4%) | 20 (19.4%) |  |
| Sometimes | 19 (22.4%) | 30 (29.1%) |  |
| Daily | 64 (75.3%) | 53 (51.5%) |  |
| **Soap use** |  |  | >0.9 |
| Never | 0 (0%) | 0 (0%) |  |
| Sometimes | 5 (5.9%) | 6 (5.8%) |  |
| Always | 80 (94.1%) | 97 (94.2%) |  |
| **Bedding washing frequency** |  |  | 0.008 |
| Rarely | 17 (20%) | 22 (21.4%) |  |
| Sometimes | 23 (27.1%) | 48 (46.6%) |  |
| Weekly | 45 (52.9%) | 33 (32.0%) |  |
| **Clothes washing frequency** |  |  | <0.001 |
| Rarely | 5 (5.9%) | 14 (13.6%) |  |
| Sometimes | 26 (30.6%) | 58 (56.3%) |  |
| Daily | 54 (63.5%) | 31 (30.1%) |  |
| **Sharing personal items** | 43 (50.6%) | 42 (40.8%) | 0.2 |
| <sup>1</sup>n (%) |  |  |  |
| <sup>2</sup>Pearson's Chi-squared test; Fisher's exact test |  |  |  |

In bivariate logistic regression, bathing frequency, clothes washing frequency, prison status, duration of incarceration, bedding washing frequency, and sharing of personal items met the inclusion criteria for multivariable analysis (Table 3).

**Table 3:** Bivariate logistic regression analysis of determinants of scabies infection.

| Variable | COR | 95% CI | p-value |
| --- | --- | --- | --- |
| **Age group** |  |  |  |
| 30–39 vs 18–29 | 0.89 | 0.46–1.73 | 0.736 |
| 40–49 vs 18–29 | 1.41 | 0.60–3.42 | 0.441 |
| ≥50 vs 18–29 | 0.51 | 0.14–1.66 | 0.271 |
| **Education** |  |  |  |
| Primary vs None | 0.27 | 0.01–1.93 | 0.252 |
| Secondary vs None | 0.27 | 0.01–1.95 | 0.256 |
| College+ vs None | 0.58 | 0.03–5.09 | 0.659 |
| **Stay >3 months** | **1.54** | **0.84–2.85** | **0.162** |
| **Remandee vs Prisoner** | **1.71** | **0.89–3.33** | **0.108** |
| **Bath frequency** |  |  |  |
| Sometimes vs Rarely | **0.16** | **0.02–0.63** | **0.021** |
| Daily vs Rarely | **0.08** | **0.01–0.30** | **0.001** |
| Soap use (Always vs Sometimes) | 1.01 | 0.28–3.47 | 0.987 |
| **Bedding washing** |  |  |  |
| Sometimes vs Rarely | 1.61 | 0.72–3.62 | 0.244 |
| Weekly vs Rarely | **0.57** | **0.26–1.23** | **0.151** |
| **Clothes washing** |  |  |  |
| Sometimes vs Rarely | 0.80 | 0.24–2.33 | 0.691 |
| Daily vs Rarely | **0.21** | **0.06–0.59** | **0.005** |
| Share personal items | **0.67** | **0.38–1.20** | **0.179** |
| Share mattress (3 vs ≤2) | 1.36 | 0.70–2.65 | 0.361 |
| Share mattress (≥4 vs ≤2) | 0.89 | 0.39–2.00 | 0.772 |
| **Ward occupancy level** |  |  |  |
| Moderate vs Low | 0.89 | 0.41–1.93 | 0.776 |
| High vs Low | 0.96 | 0.45–2.04 | 0.924 |
| Very High vs Low | 0.60 | 0.20–1.78 | 0.358 |

COR, crude odds ratio; CI, confidence interval. Reference categories are shown after "vs" for each comparison. Variables with $p < 0.20$ were included in the multivariable model.

In the adjusted individual-level model, bathing frequency and clothes washing frequency remained independently associated with scabies infection. Compared with inmates who rarely bathed, those who bathed daily had 93% lower odds of scabies infection (AOR = 0.07, 95% CI: 0.01–0.31). Similarly, inmates who washed their clothes daily had lower odds of infection than those who rarely washed their clothes (AOR = 0.21, 95% CI: 0.06–0.65). Bedding washing frequency, age, and sharing of personal items were not independently associated with infection (Table 4).

**Table 4:** Adjusted individual-level determinants of scabies infection among male inmates at Kiambu Main Prison, Kenya.

| Variable | AOR | 95% CI | p-value |
| --- | --- | --- | --- |
| **Bathing frequency** |  |  |  |
| Sometimes vs Rarely | 0.09 | 0.01–0.44 | 0.003 |
| Daily vs Rarely | 0.07 | 0.01–0.31 | <0.001 |
| **Bedding washing frequency** |  |  |  |
| Sometimes vs Rarely | 1.57 | 0.63–3.95 | 0.335 |
| Weekly vs Rarely | 0.85 | 0.34–2.10 | 0.726 |
| **Clothes washing frequency** |  |  |  |
| Sometimes vs Rarely | 0.80 | 0.23–2.54 | 0.703 |
| Daily vs Rarely | 0.21 | 0.06–0.65 | 0.007 |
| **Sharing personal items (Yes vs No)** | 0.63 | 0.32–1.25 | 0.188 |
| **Age group (years)** |  |  |  |
| 30–39 vs 18–29 | 0.68 | 0.31–1.47 | 0.319 |
| 40–49 vs 18–29 | 0.86 | 0.31–2.41 | 0.781 |
| ≥50 vs 18–29 | 0.34 | 0.07–1.36 | 0.126 |

**AOR**, adjusted odds ratio; **CI**, confidence interval. Estimates obtained from a multivariable logistic regression model including age group, bathing frequency, bedding washing frequency, clothes washing frequency, and sharing of personal items.

In the institutional model, remandees had nearly twice the odds of scabies infection compared with convicted prisoners (AOR = 1.97, 95% CI: 1.00–3.91). Duration of incarceration, receipt of health education, and lack of medicine were not independently associated with infection (Table 5).

**Table 5:**
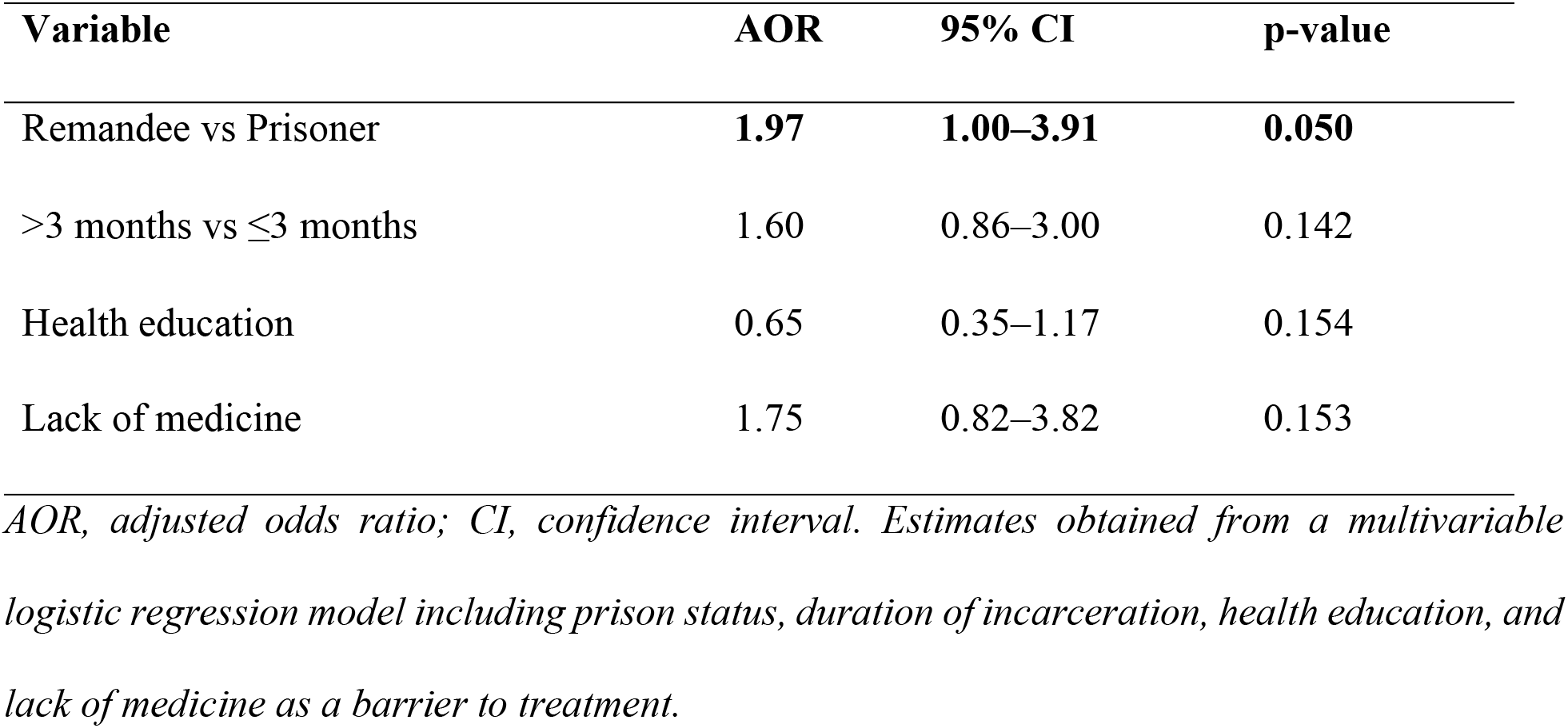
Adjusted institutional determinants of scabies infection among male inmates at Kiambu Main Prison, Kenya.

### Environmental and institutional conditions facilitating transmission

Participants identified overcrowding, poor hygiene, and admission of new inmates from police cells as key contributors to scabies transmission. Remandees were considered particularly vulnerable because they frequently arrived from police custody and often lived in more congested conditions than convicted prisoners.

“Over 100 people in a room, when they are less about 80… we sleep in a way that the legs of someone else are used as a pillow for the other.” **(FGD participant)**

“Scabies is very common, especially with new incoming inmates; they come with scabies from their respective police station cells.” **(Senior Sergeant)**

Participants also reported sharing bedding, clothing, and bathing items because of inadequate hygiene supplies, particularly soap.

“Water is always available, but soap is the problem.” **(FGD participant)**

“Most of them… depend on prison to provide essentials… if the prison does not provide them, they just stay like that, and that promotes unhygienic conditions.” **(Welfare Officer)**

### Institutional barriers to scabies prevention and management

Participants identified limited health education, delayed diagnosis and treatment, and medication shortages as barriers to scabies prevention and control.

“We have never had formal health education on scabies. They only tell us about it when someone reports having the infection; the medic just explains it casually while giving medication.” **(FGD participant)**

Participants also described delays in receiving treatment after reporting symptoms.

“A person with scabies, once they report, can take one month before getting medication, so he continues infecting others around him.” **(FGD participant)**

The clinical officer attributed these delays to financial constraints affecting medicine procurement.

“Purchasing of drugs to cater to the massive population affected is a burden for the facility.” (Clinical Officer)

## Discussion

This study found that scabies remains a major public health problem among male inmates at Kiambu Main Prison, with a weighted prevalence of 54.0%. To our knowledge, this is the first study to document the burden and determinants of scabies among incarcerated populations in Kenya. The prevalence observed was substantially higher than estimates reported among Kenyan community populations (14–16) and exceeded those reported in many prison settings globally (2,8,17–19), although it remained lower than those documented in Ghana and Uganda (20,21). The close agreement between the crude (54.8%) and weighted (54.0%) prevalence estimates indicates that the disproportionate sampling of inmates from the isolation ward had minimal influence on the overall prevalence estimate, which implies that the observed burden was representative of the wider prison population. These findings reinforce evidence that correctional facilities provide conditions that facilitate sustained scabies transmission through prolonged close contact, overcrowding, and shared living environments (2,21,22).

Personal hygiene practices emerged as important individual determinants of scabies infection. Inmates who bathed daily and washed their clothes daily had significantly lower odds of infection, consistent with previous institutional and community-based studies that identified poor hygiene as a key contributor to scabies transmission (6,7). In contrast, age, education level, soap use, and sharing of personal items were not independently associated with infection.

These findings differ from studies that reported significant associations between age, education level, and scabies infestation (6,7,21). The absence of similar associations in the present study may reflect the relatively uniform prison environment, where inmates experience similar living conditions and environmental exposures. The lack of association for soap use may be explained by the limited variability in responses, as most inmates reported using soap whenever they bathed. The lack of an independent association between sharing personal items and scabies infection is consistent with experimental evidence indicating that transmission through clothing and bedding is relatively uncommon unless such items are shared immediately after use while still warm (23). Qualitative findings, however, revealed that inadequate resources often necessitated the sharing of bedding and clothing, together with shortages of soap, detergents, and laundry facilities. The findings imply that structural conditions within prisons may have a greater influence on scabies transmission than some individual characteristics commonly associated with infection in community settings

Among the institutional factors examined, prison status appeared to be more important than duration of incarceration in explaining scabies infection. Remandees had nearly twice the odds of scabies infection compared with convicted prisoners, although the association was of borderline statistical significance after adjustment. This finding suggests that inmates in the remand section may be at greater risk of infection because of frequent admissions from police cells and the more congested conditions within remand wards, which increase opportunities for prolonged skin-to-skin contact. In contrast, the duration of incarceration was not significantly associated with scabies infection. Although inmates incarcerated for more than three months had higher odds of infection, the association was not statistically significant after adjustment. This finding differs from previous studies that reported recent admission or shorter duration of incarceration as important risk factors for scabies (8,22). The lack of association in the present study suggests that prison status may better reflect exposure to transmission than duration of incarceration, particularly in settings where remand wards experience continuous admissions and higher levels of overcrowding.

Neither the measured ward occupancy level nor the number of inmates sharing sleeping spaces was significantly associated with scabies infection in the quantitative analysis. However, qualitative findings consistently identified overcrowding as the dominant driver of transmission, with participants describing congested sleeping arrangements, close physical contact, and frequent movement of inmates from police cells as conditions that facilitated disease spread. Similar observations have been reported in prison studies across Africa, where overcrowding has been recognised as a major determinant of scabies outbreaks (2,20–22). The discrepancy between the quantitative and qualitative findings likely reflects the limited variability in ward occupancy level within Kiambu Main Prison, where most inmates were exposed to congested living conditions, reducing the ability to detect statistically significant differences.

Although receipt of health education and lack of medicine were not significantly associated with scabies infection after adjustment, both variables showed trends consistent with improved scabies control. Inmates who reported having received health education had lower odds of scabies infection, while those reporting a lack of medicine as a barrier to treatment had higher odds of infection. The qualitative findings provided contextual support for these trends, as participants reported that health education was limited, while delays in treatment were attributed to medicine shortages and financial constraints affecting drug procurement. Therefore, strengthening health education, ensuring timely diagnosis and treatment, and improving the availability of anti-scabies medication may contribute to reducing transmission within correctional facilities. In essence, these findings imply that effective scabies control in correctional facilities requires interventions that target both individual hygiene practices and structural prison conditions rather than relying on treatment alone.

### Strengths and Limitations of the Study

This is one of the first studies on scabies among inmates in Kenya and provides important baseline evidence for prison health policy and future research. The mixed-methods design strengthened interpretation by providing contextual explanations for the quantitative findings. Clinical diagnosis using the International Alliance for the Control of Scabies (IACS) diagnostic criteria and the application of sampling weights improved the reliability and representativeness of the findings. However, the cross-sectional design precludes establishing temporal or causal relationships between the identified determinants and scabies infection. Recruitment based on participant availability and willingness to participate may have introduced selection bias that could potentially limit the representativeness of the study sample. Self-reported hygiene practices may have been affected by recall or social desirability bias, which could have resulted in misclassification of some exposures. Nevertheless, the use of clinical diagnosis based on the IACS diagnostic criteria reduced the likelihood of outcome misclassification. Finally, the study was conducted in a single prison, which may limit the generalisability of the findings to other correctional facilities in Kenya or similar settings.

### Conclusion

Scabies remains a major public health concern among male inmates at Kiambu Main Prison, with more than half of the inmates affected. This study provides one of the first epidemiological assessments of scabies among incarcerated populations in Kenya and demonstrates that both individual hygiene practices and institutional conditions contribute to disease transmission. Daily bathing and daily washing of clothes were independently associated with lower odds of scabies infection, while remandees were at greater risk than convicted prisoners.

The qualitative findings provided contextual explanations for the quantitative results by identifying overcrowding, frequent admission of new inmates from police holding cells, inadequate hygiene supplies, limited health education, shortages of anti-scabies medication, and delays in diagnosis and treatment as important institutional factors that facilitate continued transmission. These findings highlight the need for integrated control measures that combine routine screening of newly admitted inmates, improved access to hygiene resources and timely treatment, strengthened prison health education, and interventions to reduce overcrowding. Further multi-centre studies are recommended to determine the burden of scabies across Kenyan correctional facilities and to evaluate the effectiveness of prison-based prevention and control strategies.

## Data Availability

The data underlying the findings of this study contain sensitive information from incarcerated individuals and cannot be shared publicly because of ethical and confidentiality considerations. De-identified data supporting the findings of this study are available upon reasonable request from the corresponding author, subject to approval by AMREF International University, the AMREF Health Africa Ethics and Scientific Review Committee (ESRC), and the Kenya Prisons Service, and completion of a data-sharing agreement to safeguard participant confidentiality.

## Acknowledgement

The authors thank the Kenya Prisons Service Headquarters for granting permission to conduct this study and the management of Kiambu Main Prison for their support. We also thank the prison staff for their assistance during data collection, particularly Ms. Nelly Chanzu for coordinating participant recruitment and conducting the clinical diagnosis of scabies. Finally, we thank all inmates who voluntarily participated in the study.

## Conflict of interest

The authors declare that there are no conflicts of interest regarding the publication of this study.

## Author Contributions

- **Conceptualization:** Esther Makokha Kulundu, Nyamongo Onkoba, Chrispine Ochieng Ngwawe.
- **Data curation:** Esther Makokha Kulundu.
- **Formal analysis:** Esther Makokha Kulundu.
- **Investigation:** Esther Makokha Kulundu.
- **Methodology:** Esther Makokha Kulundu, Nyamongo Onkoba, Chrispine Ochieng Ngwawe.
- **Project administration:** Esther Makokha Kulundu.
- **Resources:** Esther Makokha Kulundu.
- **Software:** Esther Makokha Kulundu.
- **Supervision:** Nyamongo Onkoba, Chrispine Ochieng Ngwawe.
- **Validation:** Esther Makokha Kulundu, Nyamongo Onkoba, Chrispine Ochieng Ngwawe.
- **Visualization:** Esther Makokha Kulundu.
- **Writing – original draft:** Esther Makokha Kulundu.
- **Writing – review & editing:** Esther Makokha Kulundu, Nyamongo Onkoba, Chrispine Ochieng Ngwawe.

## Supporting Information

### Supplementary Material

**S1 Questionnaire.** Structured questionnaire used to collect quantitative data from male inmates at Kiambu Main Prison.

**S2 Interview Guide.** Key informant interview and focus group discussion guides used to explore perceptions of scabies and institutional factors.

**S3 STROBE Checklist.** Completed STROBE checklist for reporting this cross-sectional mixed- methods study.

**S4 P2079-2026 Approval Letter.** AMREF ESRC approval letter.

**S5 NACOSTI RESEARCH LICENCE.** Government permit to execute the study

**S6 Kenya Prisons Approval.** Approval from the Kenya Prisons Headquarters to conduct the study

**S7 PLOS_Human_Participants_Research_Checklist_2026.** PLOS checklist

## Notes

### Competing Interest Statement

The authors have declared no competing interest.

### Author Declarations

Ethical approval for this study was obtained from the AMREF Health Africa Ethics and Scientific Review Committee (ESRC) (Approval No. P2079/2026). Research authorization was granted by the National Commission for Science, Technology and Innovation (NACOSTI). Permission to conduct the study was obtained from the Kenya Prisons Service Headquarters and the management of Kiambu Main Prison. Written informed consent was obtained from all participants prior to enrolment.

